# Surveillance and control of neglected zoonotic diseases: methodological approaches to studying Rift Valley Fever, Crimean-Congo Haemorrhagic Fever and Brucellosis at the human-livestock-wildlife interface across diverse agricultural systems in Uganda

**DOI:** 10.1101/2024.09.17.24313808

**Authors:** Dennison Kizito, Sam Tweed, Joseph Mutyaba, Nackson Babi, Swaib Lule, Gladys Nakanjako Kiggundu, Rory James Gibb, Charity Angella Nassuna, Ronald Ssali Ogwal, Ebenezer Paul, Mercy Haumba, Collins Agaba, Phionah Katushabe, Eric Morris Enyel, Stephen Balinandi, Lydia Franklinos, Naomi Fuller, Leah Owen, Laura Ferguson, Deo Birungi Ndumu, Musa Sekammatte, Atimnedi Patrick, Luke Nyakarahuka, Gladys Kalema-Zikusoka, Ibrahim Abubakar, Nigel Field, Janet Seeley, Julius Julian Lutwama, COHRIE-Uganda Team

## Abstract

**Background:** Zoonoses are of public health importance, with most major emerging diseases originating in animal populations. Rift Valley Fever (RVF), Crimean-Congo Haemorrhagic Fever (CCHF) and Brucellosis are circulating in Uganda, causing frequent outbreaks, but gaps exist in the understanding of transmission dynamics, community perspectives and effective mitigation strategies of these diseases. With increasing human-livestock-wildlife interaction in Uganda’s biodiverse cattle corridor, this study protocol outlines an integrated One Health model to determine the burden of RVF, CCHF and Brucellosis, identify key vectors and reservoirs and assesses the impact of social and policy factors on disease distribution.

**Methods:** A series of mixed-methods cross-sectional and longitudinal surveys across six conservation areas experiencing high human-livestock-wildlife interaction spanning Uganda’s Cattle Corridor: Queen Elizabeth National Park, Bwindi-Mgahinga Impenetrable Forest, Lake Mburo-Nakivaale, Murchison Falls, Kidepo Valley and Pian Upe Game Reserve. In selected villages, household surveys comprise questionnaires, focus-group discussions and in-depth interviews to determine drivers of disease risk with blood-sampling of human population. Questionnaires provide details on livestock practices, and blood sampling is conducted on cattle, sheep, pigs and goats. Targeted sampling of vectors in these localities, including mosquitoes, ticks and small mammals, using environmental traps and on-host collection. Specimens taken from nearby large wildlife include blood sampling and nasal swabs. Serological testing using indirect ELISA and molecular testing using real-time PCR was conducted to determine disease status of RVF, CCHF and Brucellosis across humans, livestock and wildlife with eco-epi modelling and qualitative analyses used to inform risks and drivers of disease.

**Results:** Baseline survey data and blood specimens were obtained from 2894 humans residing in 1602 households across 96 villages in 6 conservation areas of Uganda. A further 379 community members participated in focus group discussions and key informant interviews with 978 reformed and active poachers across 4 conservation areas. 3692 livestock were sampled, including 1925 cattle, 1409 goats, 282 sheep and 76 pigs from 358 herds. Vector data were collected for 18236 ticks, 53480 mosquitoes and 612 rodents. 241 large wildlife were sampled, including buffalo, kobs, zebras, waterbucks, topi and hartebeest. 127 Community One Health Volunteers (COHVs) were enlisted to monitor and detect outbreaks in the study sites.

**Conclusions:** This paper outlines a comprehensive One Health approach to studying neglected zoonotic diseases, integrating molecular epidemiology, social sciences and community participatory approaches involving humans, livestock, vectors and wildlife across 6 conservation areas in Uganda. It will inform interventions to enhance the surveillance and control of RVF, CCHF and Brucellosis, including strengthening outbreak preparedness and response.

## Background

### Scientific justification

Zoonotic infectious diseases are of great public importance, with most major emerging diseases originating in animals. Rift Valley Fever (RVF), Crimean-Congo Haemorrhagic Fever (CCHF) and Brucellosis are known to be circulating in Uganda, causing regular outbreaks and resulting in significant morbidity and mortality. Despite substantial human, social and economic costs, RVF, CCHF and Brucellosis have been long neglected, leading to gaps in understanding of these diseases, including strategies for mitigation and control. Further knowledge gaps include how these diseases are understood by affected communities, which practices and behaviours may contribute to driving transmission into humans, and how to best utilise One Health approaches in limiting the risk of spillover events.

### Study context

This study is conducted within and at the border of conservation areas within the cattle corridor of Uganda, where there is regular interface of domestic animals with wildlife conservation areas and forest reserves, as well as a history of the selected disease outbreaks. The region sits at a nexus of high zoonotic pathogen diversity, rapid population growth, frequent livestock movements, accelerating climatic extremes, land use changes, and substantial population dependence on ecosystems for livelihoods. These regional factors enable the study of transmission dynamics and networks of these diseases, incorporating social factors, including gendered risk variability, use of animal markets, and the impact of diversity in agricultural systems.

The Uganda Cattle Corridor covers an estimated area of 84,000km^2^ (43% of the country’s total land area) and is home to about 15 million people. The corridor is a semi-arid transition zone across the centre of the country, between the wet forest/grassland mosaics to the south on the Uganda-Tanzania border around Lake Victoria and the arid grasslands on the Uganda-Kenya-South Sudan border in the northeast (Karamoja area). The corridor is host to a mixed production system consisting of nomadic pastoralists, agro-pastoralists, and smallholder farmers. The corridor exhibits serious land and resource degradation driven by overgrazing, unsustainable agriculture practices, and charcoal production, leading to deforestation. The cattle corridor is also home to many wildlife conservation areas and forest reserves in Uganda. These factors have been noted to influence the behaviour and activities of people, vectors, and drivers of human exposure to CCHF, RVF, and Brucellosis in the country.

### Conservation Areas

The six conservation areas included cover a range of ecological and social environments, enabling understanding of specific drivers and contextual factors associated with transmission of the included zoonotic diseases:

1. Queen Elizabeth National Park is located on the equator in the Albertine Rift Valley, Western Region, Uganda. The park is contiguous with the Parc National des Virunga (Virunga National Park) in the Democratic Republic of Congo (DRC), which together completely encircle Lake Edward. The area lies at the convergence of two vegetation zones (Central African rainforest and East African grassland), with the overlap in biomes creating a range of hyper-diverse habitats supporting a vast array of birds, large carnivores and primates with human populations living in close proximity to the park.
2. Bwindi-Mgahinga Impenetrable Forest is in the far Southwest of Uganda, close to DRC, and is largely covered with the Afromontane rainforest, with the main wildlife comprising primate species, including mountain gorillas. The surrounding land is used for crop husbandry and livestock farming, with a high human population density and a vibrant international tourism industry.
3. Lake Mburo-Nakivaale conservation area lies in Southwestern Uganda and consists of primarily open grasslands, *Acacia hockii* woodlands, thickets, swamps and riverine forests surrounding Lake Mburo. It lies close to large human population settlements on the main Kampala-Mbarara highway in a region economically dependent on cattle farming with a large number of wildlife residing on public lands surrounding the park.
4. Murchison Falls conservation area is located in Albertine Rift Valley and is bisected by the Victoria Nile, comprising diverse habitats from savanna to riverine woodland and dense forest. Comprising Uganda’s largest national park, it is notable for extensive biodiversity, including over 144 mammals, 556 birds, 51 reptiles and 51 amphibian species. The Kabwoya Wildlife Reserve included in this area is a protected zone further along Lake Albert, where significant interaction with expanding human populations along the lakeshore occurs.
5. Kidepo Valley is situated in the extreme north of Uganda, bordering South Sudan and close to Kenya. It consists of dry savannah and rugged, semi-arid valleys with large numbers of lions and other game species. It represents a frontier region populated by pastoralist communities, including Karamojong, with cross-border movement of humans and animals.
6. Pian Upe Game Reserve is also located in the Karamoja sub-region in North-eastern Uganda comprising a high plateau landscape of rolling plains and wooded savanna grasslands. Settlement and grazing of domestic livestock occur within the reserve with notable levels of livestock and wildlife interaction.

### Disease background

#### RVF

Rift Valley fever virus (RVFV; *Bunyaviridae; Phlebovirus*) is transmitted to humans via contact with infected animals’ blood, body fluids, or tissues and the bites of infected mosquitoes. Over 30 species of mosquito vectors have been implicated in RVF transmission, including species from six genera: *Aedes, Culex, Anopheles, Eretmapodites, Mansonia, and Coquilletidia* [1, 2]. Floodwater *Aedes* mosquitoes are primary vectors, maintaining RVFV via transovarially infected drought-resistant eggs that can survive inter-epidemic periods. Outbreaks typically occur in 4-15-year cycles and have been consistently linked to periods of heavy rainfall and flooding, leading to semipermanent habitat for mosquitoes of other genera (e.g., secondary vectors). These vectors amplify the virus and lead to a shift from the enzootic to epizootic transmission cycle. Human infections often coincide with epizootics, primarily affecting livestock such as goats, sheep and cattle. Currently, there is no widely available human vaccine, but commercially live-attenuated and inactivated vaccines are available for livestock, and candidate human vaccines are in various stages of development and clinical testing. Uganda is known to be experiencing sporadic outbreaks in both humans and animals [3], with 27% of cattle, 7% of goats, and 4% of sheep seropositive in the Kabale district in 2016 [3] and 52 human cases of laboratory-confirmed RVF from 2017 to 2020.

#### CCHF

CCHF is a tick-borne viral zoonotic disease affecting humans, livestock and wildlife caused by the CCHF virus (CCHFV; *Bunyaviridae; Orthonairovirus*) [4]. Several tick species act as vectors for CCHF virus however, *Hyalomma* species are thought to be both the principal vector and reservoir for CCHF virus globally. The virus is maintained and transmitted in a vertical and horizontal transmission cycle involving a variety of wild and domestic vertebrate species that act as refractory hosts without showing signs of illness [5]. The ecology of CCHF in Uganda may differ from elsewhere; an investigation of a recent human CCHF case in 2015 found no evidence of the CCHF virus in Hyalomma tick species collected in the local environment. However, they did detect CCHFV in a *Rhipicephalus* (*Boophilus*) *decoloratus* tick [6]. A recent eco-epidemiological modelling study predicted that a large part of Uganda has low environmental suitability for *Hyalomma* tick species, including regions with recently confirmed CCHF cases. The study also found that the occurrence of Hyalomma was not the highest-ranked predictor of CCHF exposure, with several areas in Uganda predicted to have high CCHF exposure suitability despite low *Hyalomma* environmental suitability. This was supported by a cross-sectional study which sampled ticks from cattle at three abattoirs and found a single *Hyalomma* species tick out of a total of 1,065 ticks collected, and it tested negative for CCHF antigen [7]. CCHF antigen was detected in several ticks of the *Rhipicephalus and Amblyommma* species, again suggesting that other tick species may play an important role in CCHF virus transmission in Uganda.

#### Brucellosis

Brucellosis is a zoonotic disease caused by bacteria of the *Brucella* genus. Brucellosis is endemic in Uganda and associated with a significant disease burden, especially in pastoral communities where raw milk consumption is common [8]. Zoonotic transmission mainly occurs through direct contact with infected animals, their tissues (e.g. placenta or aborted tissues), or their products (e.g. dairy). Brucellosis infection is characterised by undulating fever and often follows a waxing and waning disease trajectory, making it difficult to diagnose and treat, resulting in many hospital visits and reduced economic activity. Brucellosis has been described in wildlife [9] and humans [8], but these studies lack critical insights from molecular epidemiology.

The viral haemorrhagic fevers (VHF) surveillance program at Uganda Virus Research Institute (UVRI), through testing samples suspected to be Ebola Virus Disease (EVD), has demonstrated that these three infections are currently circulating widely in Uganda [10]. A recent study undertaken in a high-risk group (abattoir workers) found a human CCHF seroprevalence of 12.1% (95% CI: 8.9, 16.2) among individuals reporting animal slaughtering and skinning or handling animal products compared to 9.4% (4.7, 17.7) in those with direct contact with cattle or cattle products and 4.7% (1.8, 11.8) among those tending live cattle [4]. Those with a history of tick bites were twice as likely to be seropositive (age-adjusted OR= 2.09, 95% CI: 1.09, 3.98).

### Conceptual approach and framework

One Health has received increasing prominence in research and policy priorities; however, study approaches have focused on single domains (e.g. zoonoses in humans or animals) or diseases [11]. This study adopted a novel conceptual approach, incorporating human, livestock, wildlife and vector sampling together with an examination of the social context, behaviours, laws and policies which influence their distribution. Alongside an implementation arm to establish a national community One Health surveillance programme, it aims to provide the evidence, systems and interventions to transform the landscape of One Health in Uganda.

**Figure 1.**
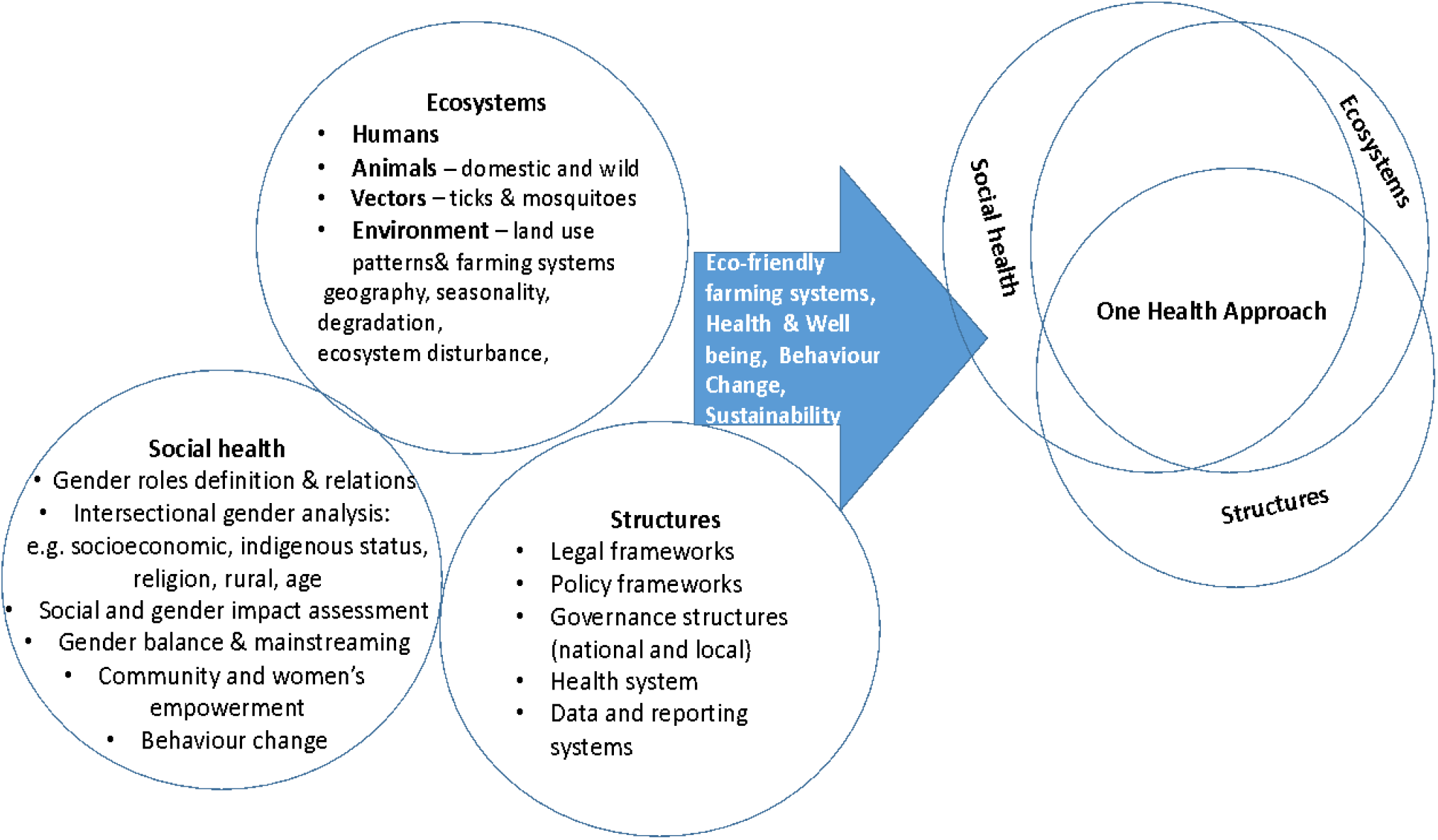
Conceptual framework underpinning consideration of social, ecological and structural determinants of disease transmission requiring One Health approach

### Aim

This study aims to improve understanding and management of RVF, CCHF, and Brucellosis using enhanced surveillance systems and a One Health approach at the human-livestock-wildlife interface in the cattle corridor of Uganda. The study will achieve this by providing burden estimations for RVF, CCHF, and Brucellosis in humans, animals, and wildlife, as well as the distribution of these infections by geography, habitat, population density, and economic activity. It will explain and inform these findings through qualitative and ecological approaches to determine behaviours influencing risk and exposure, including focused work with targeted groups, including active and reformed poachers. Management of the disease burden will be informed by vector and wildlife surveys to explain which animals act as key reservoir species for sustaining disease transmission and surveys of community leaders and policymakers to understand how national and local policies impact the level and distribution of these infections.

## Methods

A series of mixed-methods research studies collected data on transmission dynamics and networks of three zoonoses in diverse agricultural systems which interface with conservation areas within the Uganda cattle corridor. Quantitative and qualitative data collection included household surveys, in-depth interviews and focus group discussions with district technical teams and community members, alongside livestock, wildlife, vector and environmental sampling in these locations.

### Study site selection

Uganda legislatively defines conservation areas as management zones constituting national parks, forests and wildlife reserves [12]. The six areas chosen for this study represent Uganda’s largest and most prominent conservation areas, covering a diverse range of environmental zones that influence human, livestock, and wildlife interactions. All administrative districts interfacing with these conservation areas were mapped using ArcMap version 10.6. Site visits and verifications were then carried out to select districts where there was observed to be high interaction of humans, livestock and wildlife. These districts were termed Satellite Research Sites (SRS) and were limited to those inside and within the 5km radius distance from the wildlife park boundary. Up to 6 SRS were selected per conservation area and were plotted using ArcGIS software.

Within these SRS, initial mobilisation was completed by visiting each district, interfacing with the conservation area, meeting the political and local administrative leadership to introduce the project, and seeking permission to conduct the study. Subsequently, at the community level, the local council leadership was engaged and introduced to the project. During these mobilization engagements, the technical wing at the district and the sub-district level helped in identifying local resource persons to support mobilization for the project-related activities. At each conservation area, visits were conducted to each of the District Local Government offices in the respective conservation areas including the Resident District Commissioners, Chief Administrative Officers and also to inform the other key officials (District Health Officers, District Veterinary Officers, Senior Environmental Officers) about commencement of field activities. These meetings were also used to facilitate district-level key informant interviews for the qualitative study arm.

For each SRS, eligible villages were identified which had experienced changes in land cover, previous disease outbreaks, notable transboundary effects, high interface with wildlife, livestock, farming systems, and/or change in forest cover. From eligible villages, 96 were selected for sampling based on the following criteria:

1. those which interfacing with the park within a 5km radius on ArcGIS
2. those with 10km proximity of each other to facilitate efficient sampling and mobilisation and sensitisation of community members
3. those within same sub-county
4. those where supportive engagement with community leaders existed

At the village level, the ^1^Local Council 1 (LC1) was engaged before any activities were conducted. The LC1 chairperson helped to assign either an LC1 committee member or Village Health Team member as guides who introduced the research team to the respective households. At this time, the research team ensured community members were aware of the benefits of the study and the importance of their participation. The study team sought consent from each of the eligible household members who agreed to take part in the study. Prior engagement with district and village leadership enabled all identified villages to be included in the study, as no village refused participation at this stage. During the community mobilization, and other project implementation, a collaborative, multisectoral, and transdisciplinary approach of all consortium partners was always emphasized all at levels and all times.

**Figure 2.**
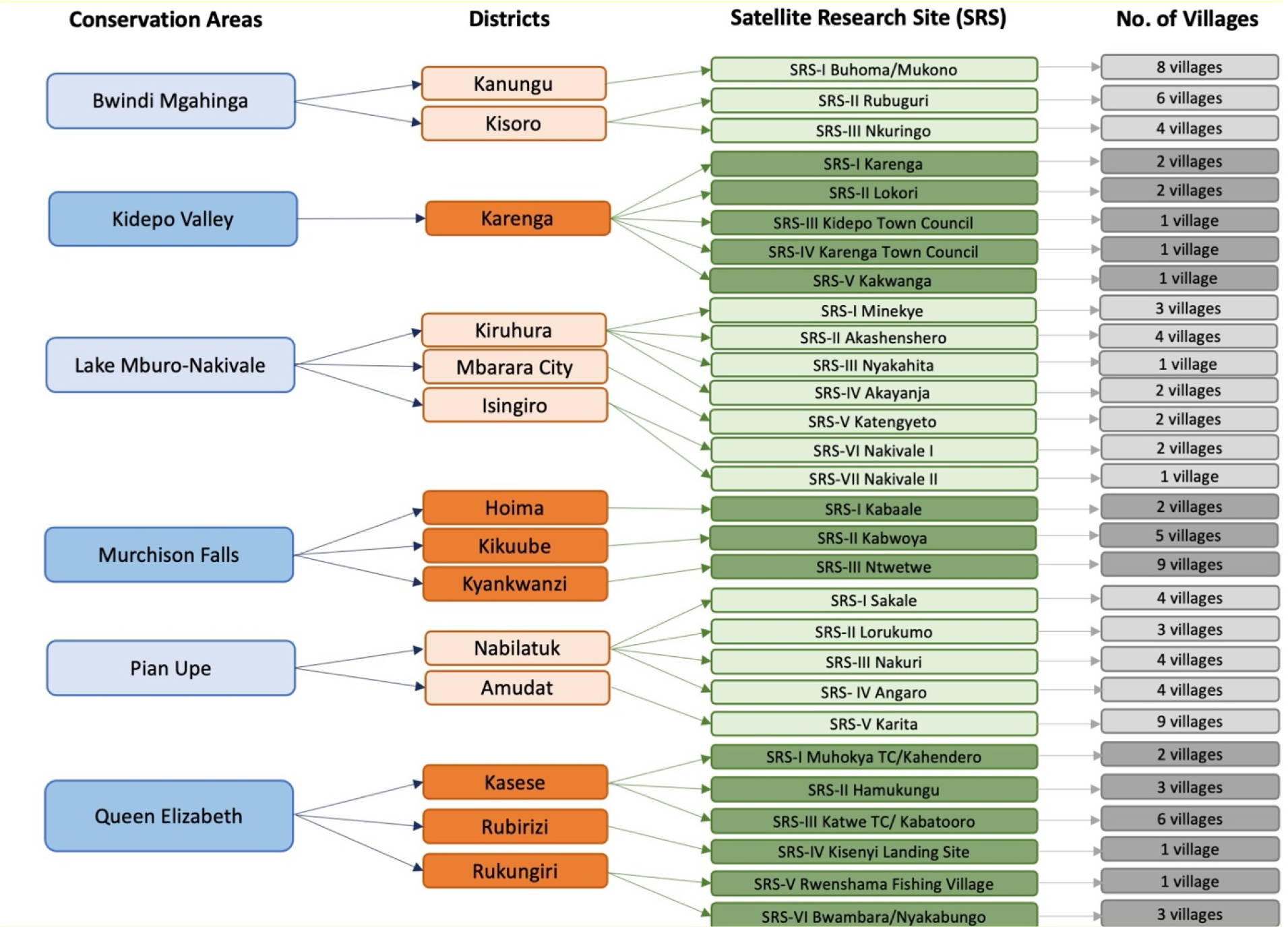
Flowchart of selected SRS and villages across six conservation areas in Uganda

**Figure 3.**
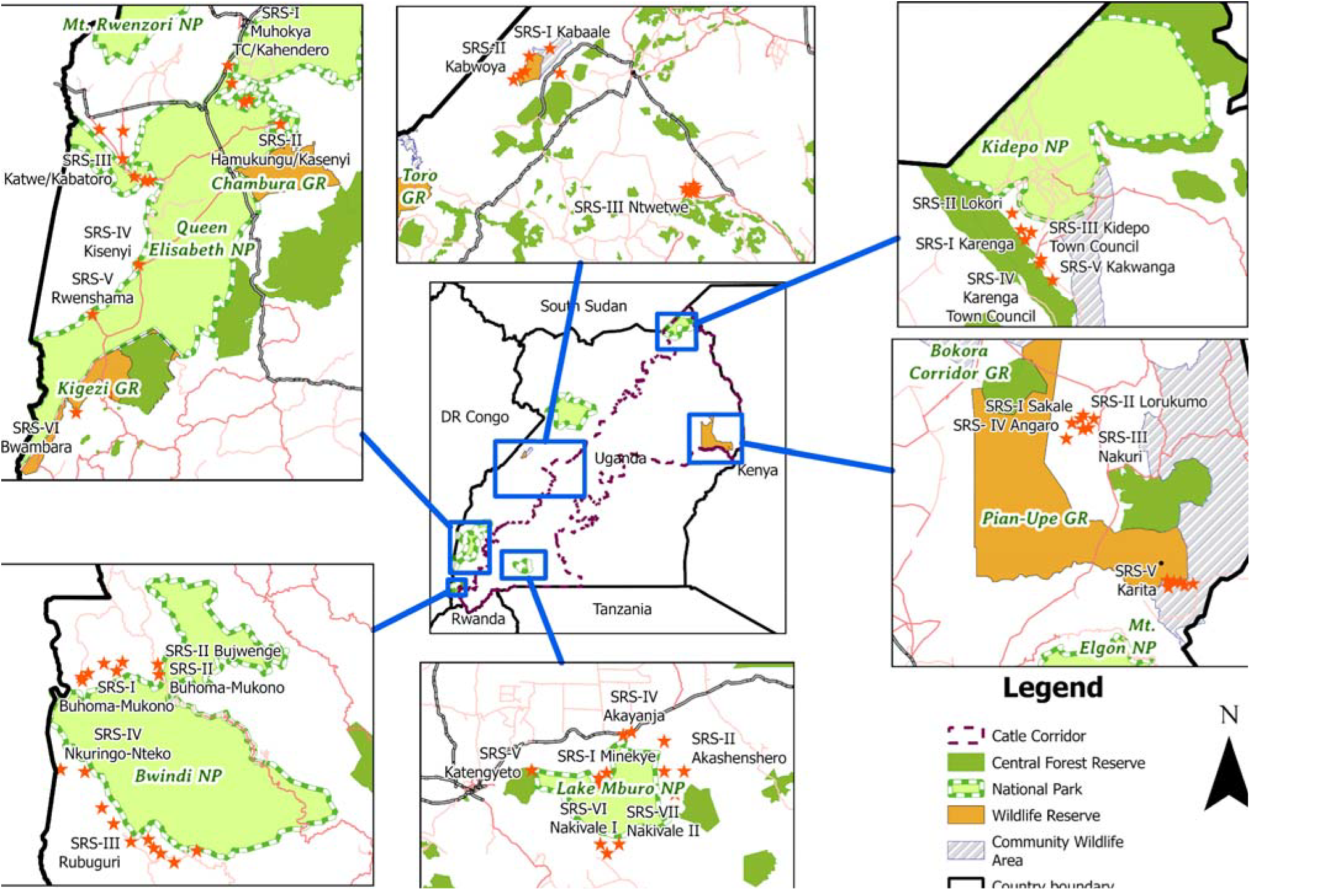
Map of selected SRS and villages (red stars) across six conservation areas in Uganda ©COHRIE-Uganda

### Survey methodology

A cross-sequential study design has been used to gather data from humans, animals (both domestic and wild), and vectors in conservation areas bordering six national parks/wildlife reserves in Uganda. Sampling has occurred during both dry and wet seasons to capture seasonal variation, disease transmission networks, and associated human activities in these areas. Data were entered on mobile tablet devices using a bespoke data collection KoboCollect^©^ app.

Household surveys were conducted to assess the knowledge, attitudes, and practices related to but not limited to the following aspects: individual socio-demographics, farming, and gender-related activities, human-animal-environment l interactions, disease prevention options, water, sanitation, and hygiene (WASH) practices, use of protective gear, nutrition and feeding habits, knowledge and risk perception related to disease (CCHF, RVF and Brucellosis) transmission, and access to and utilization of health care services.

In-depth interviews were carried out with district technical staff such as the District Health Officers, District Veterinar Officers and Senior Environmental Officers to understand the extent to which relevant laws and policies have been implemented and the impact this has had. The key informant interviews and focus group discussions were also used to collect data on local understandings of the three diseases, beliefs on root drivers, local constructions of gender and other social inequalities, socioeconomic drivers of high-risk activities, behavioural disease drivers, and indicators of community resilience.

### Human surveys

Human surveys have been conducted in adults around Bwindi Mgahinga, Kidepo Valley, Lake Mburo-Nakivale, Murchison Falls, Pian Upe and Queen Elizabeth conservation areas. Written informed consent to participate was obtained as per local ethics approval from fully trained, culturally, and linguistically competent staff from UVRI.

#### Household surveys

The selection of households for sampling within villages was based on the following criteria:

1. Proximity of households within the village to facilitate efficient movement of data collection teams
2. Balance of houses on the main road routes out of the village and remote from the village centre

Villages were scanned with the assistance of the LC1 chairperson with routes through villages mapped and all households within the village mapped. Each household was assigned a number and electronic random selection was applied to select household numbers. Where mobilisation team visited selected households and household members were not available for recruitment neighbouring households were approached instead. All members of selected households were identified with help of LC1 chairperson, including children aged 6-18 years. Children aged 5 years and under were excluded from the study due to logistical constraints of study team. Sample size calculations were based on an estimation of 13 households per village with average of 4 members per household.

Each household was requested to complete a questionnaire on occupational hazards, behaviours and cultural practices which may influence exposure risk to zoonotic infections and provide a blood sample. Household visits took around 30 minutes inclusive of signing the consent forms and blood sample collection for each participant.

Each household survey included data on the following:

- individual socio-demographics
- farming practices and household roles including consideration of gender
- human–animal-environmental interactions
- disease prevention activities (including use of mosquito nets)
- water, sanitation and hygiene practices
- use of protective gear at work, nutrition and feeding habits
- knowledge and risk perception related to disease (CCHF, RVF and Brucellosis) transmission
- access to and utilisation of health care services

For each participant a blood sample was provided which was subsequently tested for CCHF, RVF and Brucellosis by UVRI and results relayed back to the individuals alongside counselling/signposting to services and reporting to relevant public health authorities.

#### Qualitative interviews

In addition to household surveys, a series of in-depth interviews were carried out in the same communities with district technical officers including District Health Officers, District Veterinary Officers and Senior Environmental Officers. The purpose of these interviews was to understand the extent to which relevant laws and policies have been implemented and the impact this has had alongside their perspective on the CCHF, RVF and Brucellosis transmission dynamics. The In-depth interviews (IDI) and focus group discussions (FGD) also collected data on local understandings of the three diseases, beliefs on root drivers, local constructions of gender and other social inequalities, socioeconomic drivers of high-risk activities, behavioural disease drivers and indicators of community resilience. The interviews lasted between 45 minutes to 1 hour for IDIs, and 1.5 hours for FGDs.

#### Poaching community surveys

Additional in-depth interviews, household surveys and sample collection were conducted with active and reformed poachers in 4 conservation areas (Bwindi-Mgahinga, Queen Elizabeth, Lake Mburo and Murchison Falls) to further assess drivers of human-wildlife interaction. In Bwindi-Mgahinga and Queen Elizabeth conservation areas there were existing groups of reformed poachers who were approached, mobilized and invited for interview along with their household members. In Lake Mburo and Murchison Falls conservation areas, district and local leaders assisted in identifying residents in their area who were active poachers.

Specific questionnaires were administered to reformed and active poachers, focussing on assessing their knowledge of zoonotic diseases, animal contact history, current contact with wildlife, cultural practices and beliefs. Informed verbal consent was sought for the interview and for obtaining a blood sample from each household member in households where there were active or reformed poachers. We could not obtain written informed consent from participants due to safety concerns and because poaching is illegal in Uganda. Some individuals who agreed to interview, subsequently declined to give blood sample. In addition to household visits, focus group discussions were held with selected community members. In-depth interviews were conducted with group leaders of reformed poacher organisations and selected members of community where poaching was noted to be occurring.

### Livestock surveys

Survey data was collected on cattle, goats, pigs, and sheep across all 6 conservation areas. The same selection criteria were used for selection of villages as for household surveys, including areas of high human-livestock-wildlife interaction. Written informed consent to participate was obtained from herd owners in relevant areas. Recordings are made of the animal’s characteristics including presence of tick species, which are then sampled. A blood sample is provided from each surveyed animal which is tested for CCHF, RVF and Brucellosis by UVRI.

The cross-sectional study surveys across all the conservation areas employed a multi-stage sampling of Satellite Research Sites and farms within the SRS, which involved.

1. Blood sampling of livestock randomly at herd level following the consent from the farmer.
2. Collection of on-host ticks from the livestock that have been randomly sampled.

The target population of domestic ruminants (cattle, goat and sheep) and pigs in each study area calculated following the formula by Thrushfield (2018) [13] was 768. The livestock owners were mobilised beforehand by Village Health Teams and extension workers regarding the sample collection of their animals. Prior to the collection of specimens, an explanation about the background of the research activity was given by the Research Assistant. The herd inclusion/exclusion criteria of sample collection were based on;

1. Location of a herd in an area within a 5km radius to the national park or game reserve.
2. Location of a herd in an area with a history of zoonotic disease outbreaks in and around these conservation areas.
3. Natural resources used by herds like lakes and rivers are shared by all the populations that are of interest in this research project
4. Use of communal grazing land where both domestic and wildlife interact

Serum and whole blood were collected using Serum Separating Tubes and EDTA tubes respectively, from either the jugular vein or the tail vein following proper restraint from a crush or an animal handler. The samples were collected by a laboratory technician or a veterinarian. The collected samples were uniquely labelled and stored under ice awaiting transportation to the nearby veterinary laboratory for aliquoting and storage. Ticks were hand-picked across different parts of the same animals used for blood sampling by an entomologist from UVRI assisted by the district entomologists. The tick samples were placed in a falcon tube and given the same unique identification code as the blood samples collected from the animal. All the data was captured using District Health Information System Version 2 (DHIS2) and ArcGIS survey 123 software applications. Some of the electronic data that was collected during this survey were location data, details of the herd owner, the following herd variables; frequency of park grazing, herd size, vaccination status of the animals, acaricide use on the farm, watering method of the animals, grazing pattern used in the herd. Individual animal variables including age, sex, breed, origin of the animal were recorded with a unique identification given to the sample collected from that particular animal.

### Wildlife surveys

Wildlife surveys were conducted in all six conservation areas. The location of wildlife sampling was based on the same selection criteria as for household sampling.

#### Small mammal sampling methods

Small mammals present a transmission route for zoonotic disease, through both direct (bite, contact), indirect (urine, faeces) means, through vectors (ticks, fleas, mosquitoes, mites) or when predated by other species. Blood samples and any ticks present are taken from small mammal species across settings including domestic, peridomestic and sylvatic habitats and then tested for CCHF, RVF and Brucellosis by UVRI.

Small mammals were sampled inside houses (domestic), areas around the houses (peri-domestic), and extended in areas away from homes (sylvatic). Trapping sites were selected and aggregated into clusters based on. Live small animal traps were set inside houses (domestic), close to homesteads (peridomestic), beyond the homestead boundary and up to the edge of the national park (i.e. wildlife zone) targeting grasslands, bushes, and forests where gardens existed between the settlements. Homesteads were systematically selected in the villages with the help of the VHTs and other leaders starting from the village centre and following the cardinal direction skipping one house. The traps were set in the evening and checked in the morning between 8:30 and 9:30 AM. All the data on a captured animal was entered electronically using District Health Information System Version 2 (DHIS2) and ArcGIS survey 123 software applications. Data collected during this survey included location, habitats and species of the captured animals. Live trapping was adopted to get blood samples from the animals alongside a collection of ecto-parasites. Targeting the homesteads and the surrounding areas around the homesteads enabled assessment of small animal species close to people’s residences to assess which rodent and shrew species pose a risk for zoonoses transmission.

Live small animal trapping was carried out using Sherman and Tomahawk traps with the use of baits including peanut butter, silverfish, maize flour, banana and sweet potato. The captured animals were sedated using halothane, their morphometric features such as body, tail, ear and hind right foot length and the body weight were taken to help in the animal identification. The blood samples were collected by heart puncture. The blood samples were collected on DBS for easy storage and preservation. Ectoparasites were checked by combing the small animal fur and samples collected were preserved in a combination of ethanol (70%), formaldehyde (1 %) and glycerol (5%) mixture. This mixture enabled keeping the ecto-parasites in their normal colour to facilitate speciation.

#### Large wildlife sampling methods

Large wildlife were sampled to examine potential reservoirs for the CCHF, RVF and Brucellosis and determine their role in transmission across wildlife-livestock-human interactions. Fully trained and equipped staff from the Uganda Wildlife Authority perform all interactions with large wild animals, which include buffalo (*Syncerus caffer*), Uganda kob (*Kobus kob thomasi*), waterbuck (*Kobus ellipsiprymnus*), plains zebra (*Equus quagga*), hartebeest (*Alcelaphus buselaphus*) and topi (*Damaliscus lunatus jimela*). Samples are taken including blood, nasal swabs and ticks, which are then tested for CCHF, RVF and Brucellosis by UVRI.

Large wildlife samplings were conducted in five conservation areas: Queen Elizabeth, Lake Mburo, Murchison Falls, Bwindi-Mgahinga and Kidepo Valley. Sampling sites were selected as areas with intense human-wildlife-livestock interactions and selection of the species was opportunistic depending on the wildlife present in each site during the visit. Care was taken not to sample young or vulnerable animals including any animals visibly pregnant. Selection was targeted on wildlife with typical body condition, avoiding animals which appeared weak or at risk from darting. The sample size in each conservation area varied due to the availability of wildlife close to satellite research sites, climatic conditions including rainfall and the difficulty of accessing terrain by road.

Large wildlife were captured using darting with a combination of opioids and sedatives to enable sample collection. Wildlife herds were carefully approached by car and candidate animals for immobilization selected. Lone individuals were equally approached in the same way. Once identified, the prepared dart was loaded into a C02 gas powered Dan-inject JM2 dart rifle. The pressure of the dart rifle was appropriately adjusted based on the approximate distance to the animal, terrain, wind speed/direction and vegetation types but averaged 5-12 bars for a distance of 15-40 meters. The dart was delivered remotely into the candidate animals targeting large muscle mass of the shoulders, neck and thigh and animals were fully immobilized between 5-12 minutes. Immobilized animals were then approached, blindfolded, placed in sternal recumbency and stabilized. The physiological parameters were noted, and samples extracted. Doxapram was administered to enhance respiration where the need arose. Animals exhibiting high temperatures were cooled using water poured on their body. Care was taken not to work during the extreme heat of the sun.

Once the operations were completed, the rest of the team members moved to safe distance and veterinarians revived the animals. On reversal, the animals regained consciousness, got up and ran away within 1-2 minutes. The process was modified for baboons as they have a higher level of human interaction so required a combination of physical and chemical capture methods. The physical method involved the local construction of cages measuring 1.5m long x 1m wide x 1m high using local materials/poles tied with strings with a trigger-drop door as a trap with ripe bananas used as bait. The capture team waited in a hide and regularly checked the cages at intervals. In case baboons were trapped, they would have been chemically immobilized with Ketamine Hcl (250mg/ml) using a blow dart and samples as earlier described.

### Vector surveys

Mosquitoes and ticks were sampled across all six conservation areas then subsequently identified to species level by use of taxonomical keys. The primary purpose of obtaining information and collecting vectors was to determine the geographical and temporal distribution and phylogenetic focus on targeted host species alongside presence of CCHF, RVF and Brucellosis pathogens. Both cross-sectional and longitudinal surveys were carried out. Cross-sectional survey activities assessed the abundance and distribution of vector species. Longitudinal surveys involved sampling of the same sites during different seasons to determine population dynamics.

#### Mosquito sampling methods

Both indoor (resting and flying mosquitoes) and outdoor (adults, gravid, larvae, and ova/pupae mosquitoes) collections were carried out. Households were randomly selected within the study villages and indoor mosquitoes were selected using domestically recommended and approved pyrethrum spray methods and or aspirators following study-approved Indoor Standard Operating Procedures. For the outside mosquitoes CDC light traps (Miniature) were used to capture adult mosquitoes. Gravid traps were used capture gravidae and egg-laying female mosquitoes. Ovi-traps or Sticky traps for ova and pupae Larvae depending on the size and type of breeding site, larvae were collected by netting, dipping or aspirating [14, 15]. Field sorting of mosquito pools was done in batches of 50 placed in 2mls storage vials, identified by trap date, GIS location, conservation area, SRS, and village, and then transported on dry ice to One Health Laboratory (OH-Lab) at Uganda Virus Research Institute (UVRI) in Entebbe Uganda. Laboratory identification of mosquitoes was done at OH-UVRI entomology labs according to species, sex, gravidity, and blood meal presence (engorged and depleted) and identified using available keys [16] and then pooled in batches of 25 mosquitoes per species in vials with a storage medium and stored in −80^0^C freezers for further laboratory analyses [17].

#### On-host tick sampling methods

On-host tick collection was carried out from livestock, wildlife, and rodents that were sampled during the various studies. For livestock and wildlife, ticks were handpicked from various predilection sites on the body of the animal, on the face, at the horn bases, below the tail, around the udder, scrotum, penile sheath, and at the junction of the base of hooves. They were collected into labelled falcon tubes that had a similar identification as the animal that was sampled. Information regarding presence or absence of ticks on the sampled animal was entered into ArcGIS Survey 123 software application. For rodents, the ticks were collected by combing through the animal fur and placed in well labelled falcon tubes with 70% ethanol, 1%formaldehyde, and 5% glycerol mixture for preservation. Any data regarding collected ecto-parasites was entered into ArcGIS Survey 123 software application.

#### COHVs

127 Community One Health Volunteers (COHVs) were enlisted to monitor and detect outbreaks in the study sites. These volunteers include village health teams, community conservation/scouts (UWA), animal husbandry officers and disease surveillance from livestock sectors and received training on disease reporting. Regular surveillance reports were provided on mobile tablet devices with the information delivered to the relevant authorities for local response activities.

### Environmental data

#### In-situ data collection

An ecological exploration checklist was developed and used in two study areas – Pian Upe and Murchison Falls. The checklist sought to capture data that could be used as project baseline regarding ecology and environmental characterization of the study sites. Inquiries on general observation in terms of terrestrial or aquatic environments were. Major land use types were recorded depending on whether a site was forest, crop, cropland and native/natural vegetation mosaics, scrub or grassland among terrestrial habitats. Aquatic habitats were broadly checked for whether they are flowing, non-flowing or wetlands and water bodies. During the household mapping using the Open Data Kit (ODK) the following information among others was obtained on the crops grown in the area, livestock kept, dominant land use and land cover change.

#### Remote sensing variables

Climatic data in the form of satellite imagery (earth observation data source) for rainfall and temperatures were obtained from University of California, Santa Barbara. Monthly Average values for rainfall in millimetres and temperatures in degrees centigrade were extracted by sub county in QGIS version 3.28 for all satellite research sites and can be aggregated to district up to conservation area. The choice of CHIRPS rainfall is due to them being easily accessed from sites and convenience of validation with various data sources. Rainfall data considered is from 1981 to 2024 is compiled by Climate Hazards InfraRed Precipitation with Station data or CHIRPS while that temperature was from Moderate Resolution Imaging Spectroradiometer or MODIS and enhanced Visible Infrared Imager/Radiometer Suite or eVIIRS satellite sensors for the period 2002 to 2024.

Land cover and land use change (LCLUC) were derived from national datasets generated and provided by the National Forestry Authority (NFA) by mandate. Datasets considered in the study include epochs for 1990s, 2000, 2005, 2010, 2015, 2017 and 2019. From among these, decadal variations could be assessed for each conservation area. The datasets were clipped to fit areas of interest where the research was taking place.

## Analysis Methodology

### Laboratory analyses

#### Serological analysis

An inhouse indirect ELISA was developed to detect antibodies against CCHF, RVF and Brucellosis in humans, livestock (cattle, goats, sheep, pigs), small mammals (rodents, shrews) and wildlife (kob, zebra, waterbuck, buffalo). This was completed using a checkerboard titration technique with samples obtained from the field sites and reference sera. Serial dilutions were used to determine the optimal coating antigen concentration sample dilutions and anti-species Horseradish peroxidase (HRP) conjugate where the concentration at which the highest signal-to-noise ratio was obtained was considered as optimal. The optical density value was obtained at 450nm wavelength. The cut-off values were obtained from the negative samples obtained and normal reference sera where the formula used was cut-off values = Mean OD Negative + 3SD.

Coating antigens for the in-house ELISAs included the CCHF whole antigen obtained in kind from the laboratory of Dr. Thomas Ksiazek at the University of Texas Medical Branch at Galveston, d/b/a UTMB Health. For Brucellosis we used the Recombinant B. abortus Outer Membrane Protein and for Rift Valley Fever we used recombinant nucleoproteins and conjugates from CD Creative Diagnostics, NY, USA. We did a comparability analysis using certified commercial kits manufactured by Innovative Diagnostics Grabels, France, and these included ID Screen Rift Valley Fever Competition Multi-species, ID Screen CCHF Double Antigen Multi-species, and ID Screen Brucellosis Serum Indirect Multi-species.

#### Molecular analysis

Given the large number of samples and the diversity in species collected, for nucleic acid extraction pools of 6 whole-blood samples each were made per species, a total of 1,950 pools were generated for extraction. Magmax Express 96 automated extractor was used to isolate nucleic material from all the samples using the Magmax Pathogen RNA/DNA Kit (Thermo Fisher Scientific, Carlsbad, CA) following manufacturer’s instructions.

Realtime PCR was then conducted with the probe and primer sequences used for RVF and CCHF PCR assays follow those outlined by Pang et al [18]. The primer and probe sequences used for *Brucella* spp, *Brucella abortus* and *Brucella melitensis* followed methodology published by Probert et al.,[19] whereas *B. suis, B. ovis, B. canis* and *B. neotomae* were obtained from methods outlined by the Hinić et al [20]. approach. All primers and probes were manufactured by Macrogen Europe B.V (supplementary data in annex). Probe-based one-step real time PCR assays for CCHF, RVFV and Brucellosis were set up using the qScript XLT 1-Step RT-qPCR ToughMix kit (Quantabio Beverly, MA). For Brucellosis, the first step involved screening of the DNA sample for brucellosis using the *Brucella* spp. primers and probes. The positives obtained were then subjected to a subtyping PCR using primers targeting *B. abortus, B. melitensis, B. suis, B. ovis, B. canis* and B. neotomae. All positives will be further sequenced to provide full genomic detail of the pathogens.

### Statistical analyses

Datasets including demographic descriptors, exposure data and infection status will be used to determine the burden of CCHF, RVF and Brucellosis in humans, animals, and wildlife as well as understand which wild animals act as key reservoirs for sustaining transmission. We aim to describe the socio-demographic characteristics of the study population. Furthermore, spatial modelling will be used to detail the distribution of these infections by geography, habitat, population density, and economic activity. GIS and spatial modelling will be used to map the distribution of disease cases and identify hotspots. Analyses will include ecological and epidemiological modelling including climatic and meteorological datasets to understand changing patterns of transmission across research sites.

Analyses will focus on integrating datasets taking an ecosystem perspective [10], including tick and host infection data, environmental sampling and wildlife reservoirs to answer fundamental questions on the life cycle of vectors and their relative contribution to disease transmission. The analytical approach will consider common exposures across CCHF, RVF and Brucellosis, recognising that interventions to reduce disease burden will impact multiple infection pathways. Researchers will use standard statistical programmes/languages including STATA and R for data analysis purposes.

Qualitative data analysis will involve transcribing audio recorded data into text in word. Transcripts will then be exported into NVivo 12^©^ for coding. The code book will be developed following major themes aligned to the study objectives. Any emergent themes and subthemes will be flagged and discussed and resolved by the lead social science team. Thereafter analytic memos will be utilized for preliminary data analysis with thematic content analysis used. Results will be presented as direct quotes and narratives and triangulated with quantitative results to enrich findings and provide context.

### Eco-epi modelling analyses

Statistical modelling will be conducted with inference and selection using Bayesian hierarchical regression using Integrated Nested Laplace Approximation in statistical package R [21, 22]. This framework will enable the development of spatially and temporally structured regression models that address data sparsity and spatial bias whilst also being computationally tractable [22, 23]. Subsequently binomial spatial Bayesian hierarchical models will be developed to address hypotheses including the tick infestation on individual livestock across species.

### Qualitative data analysis

Audio recorded data were transcribed, translated verbatim into English and captured in MS-Word. A team of three experienced social scientists read the transcripts and edited them for completeness. These were later uploaded into NVivo version 12 for coding. A draft code book was generated, discussed and agreed upon by a team of five social scientists to ensure the codes were aligned to the study objective. Two social scientists coded the same transcripts guided by the code book. Any new emerging themes in the data were flagged, discussed, and agreed upon before being coded. The results are presented as quotes.

### Ethics

Ethical approval was granted for this study by Uganda Virus Research Institute Research and Ethics Committee (reference GC/127/875), Uganda National Council for Science and Technology (UNCST) (number HS2053ES), Uganda Wildlife Authority (reference COD/9605), University College London (Ethics ID: 27491.001), and University of Southern California (UP-22-00138). Full study protocols were submitted in advance of mobilisation to District Authorities.

The study has obtained all necessary local permission for all aspects of the research with all interactions with study participants conducted by staff from Uganda Virus Research Institute with full training in research methods and ethical considerations, as well as being culturally and linguistically competent for interactions with populations concerned. Field teams are briefed on informed consent and during surveys, written consent is obtained from participants before sample collections, carrying out interviews, or performing any other investigations. Humans who seek withdrawal from the study have their data removed from the database and will not be included in the datasets.

Any humans found to have current infections are contacted, informed and supported. This activity is conducted by the local clinical research and social counsellors’ team who will carry out preliminary counselling and basic management before referring (if appropriate) to the nearest medical facility or hospital for definitive management and support. The herd owner(s) are informed, provided with relevant information, and directed to seek veterinary services for animals found to have infections. The research team ensured full consideration throughout the study of animal welfare, rights, and protections in line with Uganda Ministry of Agriculture, Animal Industries and Fisheries (MAAIF) guidance. All protocols for interactions with wildlife and/or livestock are agreed in advance with the Uganda Wildlife Authority and local District Veterinary authorities.

## Results

Baseline survey data and blood samples were obtained from 2894 residents residing in 1602 households across 6 conservation areas.

**Table 1:** Baseline characteristics of human participants in household surveys sampled across conservation areas.

| Baseline household human survey | No. of participants (n=2894) | % |
| --- | --- | --- |
| **Conservation Area** |  |  |
| Bwindi Mgahinga | 537 | 18.5% |
| Kidepo Valley | 205 | 6.74% |
| Lake Mburo-Nakivale | 503 | 17.4% |
| Murchison Falls | 355 | 12.2% |
| Queen Elizabeth | 604 | 20.8% |
| Pian Upe | 690 | 22.7% |
| **Age group (years)** |  |  |
| <20 | 357 | 14.1% |
| 20-29 | 1,240 | 49.0% |
| 30-39 | 368 | 14.5% |
| 40-49 | 241 | 9.5% |
| >=50 | 327 | 12.9% |
| **Sex** |  |  |
| Female | 1,815 | 59.9% |
| Male | 1,214 | 40.1% |

Community qualitative surveys were conducted with 379 individuals across 6 conservation areas including 35 focus group discussions and 67 key informant interviews.

**Table 2:** Baseline characteristics of humans in community qualitative surveys.

| Community qualitative surveys | No. of Focus Group Discussions (n=35) | No. of Focus Group participants (n=312) | No. of In-depth Interviewees (n=67) | Total No. of participants (n=379) |
| --- | --- | --- | --- | --- |

| Conservation Area |  | Male | Female | Male | Female |  |
| --- | --- | --- | --- | --- | --- | --- |
| Bwindi Mgahinga | 6 | 27 | 35 | 4 | 1 | 67 |
| Kidepo Valley | 6 | 24 | 24 | 3 | 0 | 51 |
| Lake Mburo-Nakivale | 5 | 13 | 20 | 28 | 6 | 67 |
| Murchison Falls | 8 | 32 | 28 | 6 | 2 | 68 |
| Pian Upe | 5 | 20 | 32 | 8 | 1 | 61 |
| Queen Elizabeth | 5 | 28 | 29 | 8 | 1 | 65 |

Targeted engagement was conducted with communities where poaching occurs from 4 conservation areas. This included 6 individual interviews with reformed poachers and 5 interviews with active poachers. A further 6 focus group discussions were conducted with reformed poachers and 5 focus group discussions with active poachers.

**Table 3:** Baseline characteristics of humans in targeted poaching community survey (quantitative & qualitative)

| Poaching communities | No. of participants (n=978) | % |
| --- | --- | --- |
| **Qualitative survey and specimen collections** |  |  |
| Bwindi Mgahinga | 214 | 21.9% |
| Lake Mburo-Nakivale | 226 | 23.1% |
| Murchison Falls | 282 | 28.8% |
| Queen Elizabeth | 256 | 26.2% |

A comprehensive collection of 3692 livestock, including cattle, goats, sheep and pigs from 358 herds was conducted.

**Table 4:** Baseline characteristics of livestock sampled across conservation areas.

| Livestock | No. of animals (n=3237) | % |
| --- | --- | --- |
| **Conservation Area** |  |  |
| Bwindi Mgahinga | 533 | 16.5% |
| Kidepo Valley | 394 | 12.2% |
| Lake Mburo-Nakivale | 727 | 22.5% |
| Murchison Falls | 200 | 6.2% |
| Pian Upe | 682 | 21.1% |
| Queen Elizabeth | 701 | 21.7% |
| **Species** |  |  |
| Cattle | 1925 | 52.1% |
| Goats | 1409 | 38.2% |
| Sheep | 282 | 7.6% |
| Pigs | 76 | 2.1% |
| **Age of animal** |  |  |
| <5 years old | 2831 | 87.5% |
| >= 5 years old | 386 | 12.5% |
| **Sex of animal** |  |  |
| Female | 2654 | 81.1% |
| Male | 575 | 18.9% |
| **Herd Size** |  |  |
| <50 animals | 1386 | 42.8% |
| 50-99 animals | 129 | 3.99% |
| >=100 animals | 1722 | 53.2% |

Specimens were collected from 241 wildlife across 6 conservation areas including Buffalo, Uganda Kob, Zebra, Hartebeest, Waterbuck, vectors (5838 ticks and 53480 mosquitoes) and 700 rodents (including rats, mongoose)

**Table 5:** Baseline characteristics of wildlife sampled across conservation areas.

| Wildlife | No. of animals (n=233) | % |
| --- | --- | --- |
| **Conservation Area** |  |  |
| Kidepo Valley | 45 | 19.3 |
| Lake Mburo-Nakivale | 58 | 24.9 |
| Murchison Falls | 79 | 33.9 |
| Queen Elizabeth | 51 | 21.9 |
| **Species** |  |  |
| Buffalo | 80 | 34.3 |
| Hartebeest | 3 | 1.29 |
| Waterbuck | 13 | 5.58 |
| Uganda Kob | 88 | 37.8 |
| Zebra | 48 | 20.6 |
| Topi | 1 | 0.4 |
| **Sex of animal** |  |  |
| Female | 124 | 53.2 |
| Male | 109 | 46.8 |

**Table 6:** Baseline characteristics of small mammals sampled across conservation areas.

| Small mammals | No. of animals (n=612) | % |
| --- | --- | --- |
| **Conservation Area** |  |  |
| Bwindi Mgahinga | 127 | 20.8 |
| Kidepo Valley | 87 | 14.2 |
| Lake Mburo-Nakivale | 67 | 10.9 |
| Murchison Falls | 22 | 3.6 |
| Queen Elizabeth | 309 | 50.5 |
| **Species** |  |  |
| *Acomys caharinus* | 2 | 0.3 |
| *Aethomys spp* | 2 | 0.3 |
| *Apodemus sylvaticus* | 3 | 0.5 |
| *Arvicanthus niloticus* | 87 | 14.2 |
| *Crocidura oliveri* | 5 | 0.8 |
| *Crocidura spp* | 57 | 9.3 |
| *Deomys ferugineus* | 2 | 0.3 |
| *Dephomys defua* | 2 | 0.3 |
| *Gerbilliscus spp* | 12 | 2.0 |
| *Graphiurus spp* | 4 | 0.7 |
| *Herpestes flavescens* | 1 | 0.2 |
| *Lemniscomys striatus* | 25 | 4.1 |
| *Lophuromys spp* | 31 | 5.1 |
| *Mastomys natalensis* | 89 | 14.5 |
| *Mungos mungo* | 1 | 0.2 |
| *Mus domestica* | 3 | 0.5 |
| *Mus minutoides* | 18 | 2.9 |
| *Oenomys hypoxanthus* | 4 | 0.7 |
| *Praomys spp* | 6 | 1.0 |
| *Rattus rattus* | 248 | 40.5 |
| **Sex of animal** |  |  |
| Female | 315 | 51.5 |
| Male | 295 | 48.2 |

In addition to small mammals, vectors were sampled across conservation areas, with 18236 ticks and 53480 mosquitoes collected in total.

**Table 7:** Baseline characteristics of ticks sampled across conservation areas.

| Vectors | No. of ticks sampled (n=18236) |
| --- | --- |
| **Conservation Area** |  |
| Bwindi Mgahinga | 1409 |
| Kidepo Valley | 1472 |
| Lake Mburo-Nakivale | 5039 |
| Murchison Falls | 4806 |
| Pian Upe | 3293 |
| Queen Elizabeth | 2217 |

## Discussion

The study is designed to provide a comprehensive and integrated methodology for determining the burden, drivers and impact of RVF, CCHF and Brucellosis across conservation areas in Uganda. The pathogens selected share common exposure pathways with livestock a key factor and typically co-exist in the same communities, yielding insights relevant for control across zoonoses. A limitation of the study is that a wider range of diseases were not included, and a potential risk is that biological and social risk factors differ across the diseases more than expected, meaning mitigation strategies are less comprehensive.

Research findings will be disseminated to the communities involved in the study through a series of feedback events in relevant regions, such as a Karamoja regional dissemination event covering Kidepo Valley and Pian Upe conservation areas. These dissemination activities will incorporate members of affected communities, community leaders, technical and administrative staff including District Health Officers, District Veterinary Officers and Senior Environmental Officers, political representatives from local, regional and national level. These events will provide an opportunity for increasing awareness of the diseases as well as mitigation strategies and building momentum for working more effectively across One Health at a local level.

Testing for current infections were not included in laboratory analyses, so dissemination of results on an individual basis was not the focus for this study. The dissemination events described above provided the method to share results at community level.

## Conclusions

This paper outlines a comprehensive One Health approach to studying neglected zoonotic diseases integrating molecular epidemiology, social sciences and community participatory approaches involving humans, livestock, vectors and wildlife across 6 conservation areas in Uganda. It will inform interventions to enhance the ongoing surveillance and control of RVF, CCHF and Brucellosis including strengthening outbreak preparedness and response.

## Data Availability

All data produced in the present study are available upon reasonable request to the authors and in line with agreed ethical protocols.

## Footnotes

1 Local Council 1 leadership is comprised of eleven village committee members namely: Chairperson, Vice Chairperson, General Secretary, Defence, Publicity, Women Affairs, Youth Affairs, Production & Environment, Disability Affairs, and Treasurer

## References

1. Bird BH, Ksiazek TG, Nichol ST, Maclachlan NJ. Rift Valley fever virus. J Am Vet Med Assoc. 2009;234(7):883–93. doi: 10.2460/javma.234.7.883. PubMed PMID: 19335238.

2. Linthicum KJ, Britch SC, Anyamba A. Rift Valley Fever: An Emerging Mosquito-Borne Disease. Annu Rev Entomol. 2016;61:395–415. doi: 10.1146/annurev-ento-010715-023819. PubMed PMID: 26982443.

3. Nyakarahuka L, de St Maurice A, Purpura L, Ervin E, Balinandi S, Tumusiime A, et al. Prevalence and risk factors of Rift Valley fever in humans and animals from Kabale district in Southwestern Uganda, 2016. PLoS Negl Trop Dis. 2018;12(5):e0006412. Epub 20180503. doi: 10.1371/journal.pntd.0006412. PubMed PMID: 29723189; PubMed Central PMCID: PMCPMC5953497.

4. Bente DA, Forrester NL, Watts DM, McAuley AJ, Whitehouse CA, Bray M. Crimean-Congo hemorrhagic fever: history, epidemiology, pathogenesis, clinical syndrome and genetic diversity. Antiviral Res. 2013;100(1):159–89. Epub 20130729. doi: 10.1016/j.antiviral.2013.07.006. PubMed PMID: 23906741.

5. Gargili A, Estrada-Pena A, Spengler JR, Lukashev A, Nuttall PA, Bente DA. The role of ticks in the maintenance and transmission of Crimean-Congo hemorrhagic fever virus: A review of published field and laboratory studies. Antiviral Res. 2017;144:93–119. Epub 20170601. doi: 10.1016/j.antiviral.2017.05.010. PubMed PMID: 28579441; PubMed Central PMCID: PMCPMC6047067.

6. Balinandi S, Patel K, Ojwang J, Kyondo J, Mulei S, Tumusiime A, et al. Investigation of an isolated case of human Crimean-Congo hemorrhagic fever in Central Uganda, 2015. Int J Infect Dis. 2018;68:88–93. Epub 20180131. doi: 10.1016/j.ijid.2018.01.013. PubMed PMID: 29382607; PubMed Central PMCID: PMCPMC5893389.

7. Lule SA, Gibb R, Kizito D, Nakanjako G, Mutyaba J, Balinandi S, et al. Widespread exposure to Crimean-Congo haemorrhagic fever in Uganda might be driven by transmission from Rhipicephalus ticks: Evidence from cross-sectional and modelling studies. J Infect. 2022;85(6):683–92. Epub 20220922. doi: 10.1016/j.jinf.2022.09.016. PubMed PMID: 36152736.

8. Tumwine G, Matovu E, Kabasa JD, Owiny DO, Majalija S. Human brucellosis: sero-prevalence and associated risk factors in agro-pastoral communities of Kiboga District, Central Uganda. BMC Public Health. 2015;15:900. Epub 20150915. doi: 10.1186/s12889-015-2242-z. PubMed PMID: 26374402; PubMed Central PMCID: PMCPMC4572625.

9. Aruho R, MacLeod ET, Manirakiza L, Rwego IB. A serological survey of brucellosis in wildlife in four major National Parks of Uganda. BMC Vet Res. 2021;17(1):95. Epub 20210301. doi: 10.1186/s12917-021-02782-4. PubMed PMID: 33648507; PubMed Central PMCID: PMCPMC7923651.

10. Shoemaker TR, Nyakarahuka L, Balinandi S, Ojwang J, Tumusiime A, Mulei S, et al. First Laboratory-Confirmed Outbreak of Human and Animal Rift Valley Fever Virus in Uganda in 48 Years. Am J Trop Med Hyg. 2019;100(3):659–71. doi: 10.4269/ajtmh.18-0732. PubMed PMID: 30675833; PubMed Central PMCID: PMCPMC6402942.

11. Gibb R, Franklinos LHV, Redding DW, Jones KE. Ecosystem perspectives are needed to manage zoonotic risks in a changing climate. BMJ. 2020;371:m3389. Epub 20201113. doi: 10.1136/bmj.m3389. PubMed PMID: 33187958; PubMed Central PMCID: PMCPMC7662085.

12. UWA. Uganda Wildlife Authority. 2022 April 30,

13. Epidemiology V. Veterinary Epidemiology 4th Edition. 4th Edition ed.

14. Kaddumukasa MA, Mutebi JP, Lutwama JJ, Masembe C, Akol AM. Mosquitoes of Zika Forest, Uganda: species composition and relative abundance. J Med Entomol. 2014;51(1):104–13. PubMed PMID: 24605459.

15. ECDC. Field sampling methods for mosquitoes, sandflies, biting midges and ticks – VectorNet project 2014–2018. Stockholm and Parma: ECDC and EFSA; 2018. Stockholm, Sweden: 2018 ISBN 978-92-9498-251-3 Contract No.: Catalogue number TQ-02-18-863-EN-N.

16. Edwards FW. Mosquitoes of the Ethiopian Region III. Culcine adults and pupae. London, United Kingdom: London British Museum of Natural History.; 1941.

17. Mossel EC, Crabtree MB, Mutebi JP, Lutwama JJ, Borland EM, Powers AM, et al. Arboviruses Isolated from Mosquitoes Collected in Uganda, 2008-2012. J Med Entomol. 2017;54(5):1403–9. doi: 10.1093/jme/tjx120. PubMed PMID: 28874015; PubMed Central PMCID: PMCPMC5968633.

18. Pang AW, Macdonald JR, Yuen RK, Hayes VM, Scherer SW. Performance of high-throughput sequencing for the discovery of genetic variation across the complete size spectrum. G3 (Bethesda). 2014;4(1):63–5. Epub 20140110. doi: 10.1534/g3.113.008797. PubMed PMID: 24192839; PubMed Central PMCID: PMCPMC3887540.

19. Probert WS, Schrader KN, Khuong NY, Bystrom SL, Graves MH. Real-time multiplex PCR assay for detection of Brucella spp., B. abortus, and B. melitensis. J Clin Microbiol. 2004;42(3):1290–3. doi: 10.1128/JCM.42.3.1290-1293.2004. PubMed PMID: 15004098; PubMed Central PMCID: PMCPMC356861.

20. Hinic V, Brodard I, Thomann A, Cvetnic Z, Makaya PV, Frey J, et al. Novel identification and differentiation of Brucella melitensis, B. abortus, B. suis, B. ovis, B. canis, and B. neotomae suitable for both conventional and real-time PCR systems. J Microbiol Methods. 2008;75(2):375–8. Epub 20080715. doi: 10.1016/j.mimet.2008.07.002. PubMed PMID: 18675856.

21. Lindgren F, Rue H. Bayesian Spatial Modelling with R-INLA. Journal of Statistical software 2015;63. doi: 10.18637/jss.v063.i19.

22. Rue H, Martino S, Chopin N. Approximate Bayesian Inference for latent Gaussian models by using intergrated nested Laplace approximations. Journal of the Royal Statistical Society: series B (Statistical Methodology). 2009;71(2). doi: 10.1111/j.1467-9868.2008.00700.x.

23. Redding DW, Tiedt S, Lo Iacono G, Bett B, Jones KE. Spatial, seasonal and climatic predictive models of Rift Valley fever disease across Africa. Philos Trans R Soc Lond B Biol Sci. 2017;372(1725). doi: 10.1098/rstb.2016.0165. PubMed PMID: 28584173

